# A Clinical Guideline-Grounded Hybrid Agentic Framework for Holistic Epilepsy Management

**DOI:** 10.64898/2026.03.17.26348205

**Authors:** Duy Khoa Pham, Dinesh Giritharan, Guilherme Camargo de Oliveira, Bao Vo Quoc, Karin Verspoor, Meng Law, Patrick Kwan, Zongyuan Ge, Deval Mehta

## Abstract

Epilepsy is a chronic neurological disorder requiring multi-faceted management, including seizure detection, syndrome diagnosis, prognostication, antiseizure medication recommendation, epileptogenic zone localization, and surgical outcome prediction. Although numerous deep learning approaches have been developed for individual tasks, these models are typically siloed and modality-specific (e.g., EEG for seizure detection, MRI for localization), failing to reflect the multidisciplinary nature of real-world epilepsy care, where epileptologists, neuroradiologists, neurosurgeons, neuropsychologists and neuropsychiatrists jointly interpret heterogeneous evidence to guide decisions. In this work, we propose a clinical guideline-grounded hybrid multi-agent framework for holistic epilepsy management. Heterogeneous patient data is processed through modality-specific discriminative and generative models, where textual interpretations from generative agents are combined with structured predictions from discriminative models as auxiliary guidance. This aggregated evidence is passed to a central orchestrating agent grounded in international epilepsy guidelines, which evaluates multi-modal findings within structured clinical pathways and performs iterative cross-agent coordination for evidence-informed decision-making. We evaluate our framework across two datasets spanning six epilepsy management tasks and also introduce a publicly available multi-modal, multi-task epilepsy benchmark. Results demonstrate that integrating discriminative evidence with guideline-grounded generative coordination yields more reliable and comprehensive decisions compared to conventional LLM-based and task-specific baselines. Our dataset and code is available at [URL](https://anonymous.4open.science/r/epi_guide-EA18).

## 1 Introduction

Epilepsy is a chronic neurological disorder characterized by recurrent seizures due to abnormal brain activity, affecting over 60 million people worldwide [18]. Notably, more than one-third of patients develop drug-resistant epilepsy, failing to achieve seizure control despite trials of multiple appropriate medications [13]. Given its lifelong and heterogeneous nature, epilepsy requires continuous, multi-stage management – from seizure and syndrome diagnosis to epileptogenic zone (EZ) localization and treatment decisions such as anti-seizure medication (ASM) selection or surgical intervention. These decisions depend on integrating heterogeneous data, including electroencephalography (EEG), magnetic resonance imaging (MRI), genomics, and diverse clinical factors, necessitating coordinated multi-modal analysis.

Numerous automated methods have been developed for individual epilepsy tasks (Fig 1-a), including seizure type classification from EEG [3] and video [16,10], lesion localization from MRI [2,23], EZ prediction from semiology reports [28], and ASM response prediction from clinical and imaging data [20]. More recently, foundation models (FMs) and large language models (LLMs) have been explored for modality-specific analysis and reasoning (Fig 1-b), such as EEG-based report generation [11], automated event detection from health records [9], and explanation-driven ASM recommendation [22]. However, despite these advances, existing methods remain siloed, limited to single tasks and specific modalities, failing to reflect the coordinated, multi-task nature of real-world epilepsy care.

**Fig. 1.**
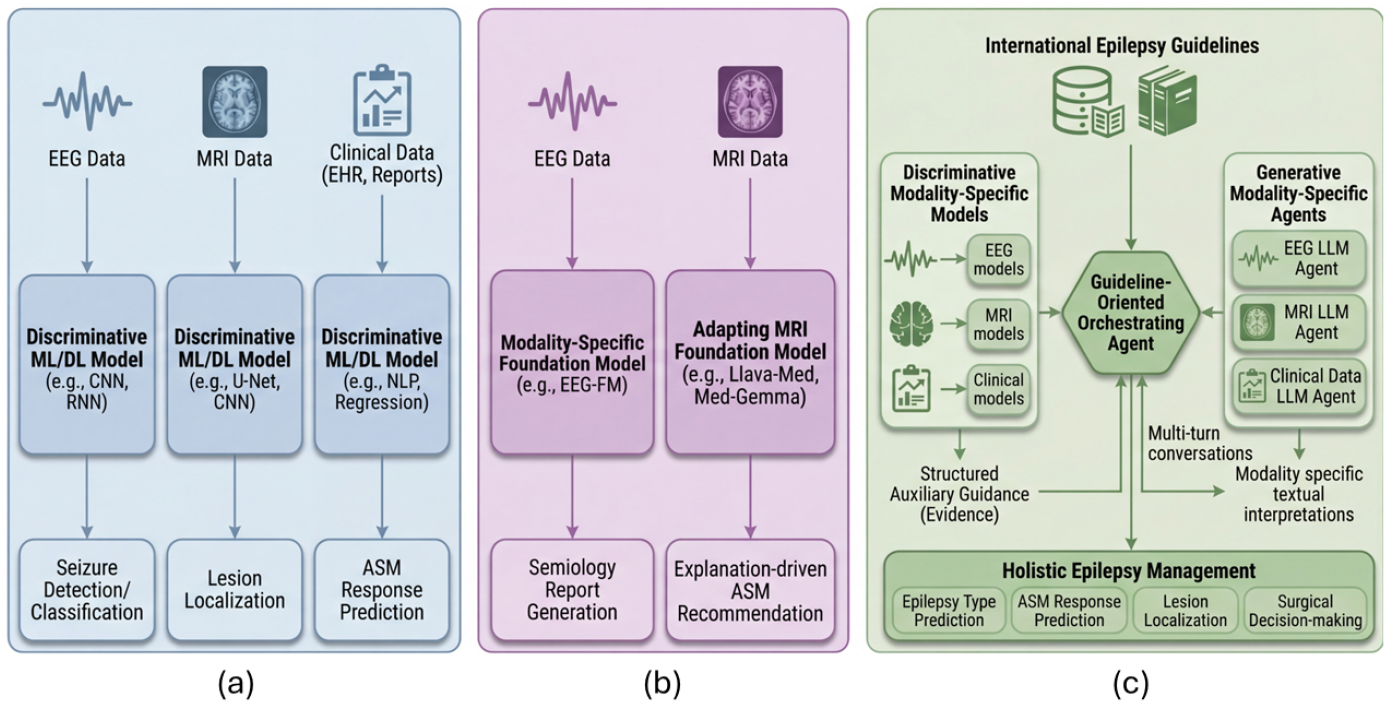
Comparative overview. (a) *Siloed discriminative models* trained on modality-specific data for individual tasks; (b) *Siloed foundation models* applied to single-task, single-modality settings; (c) *Our proposed hybrid framework* coupling discriminative models and generative agents for multi-modal, multi-task holistic epilepsy management.

More broadly, the medical AI community has seen rapid growth in generative multi-agent systems that coordinate role-specific LLMs for complex clinical tasks [6,12,25,14,4]. These frameworks decompose problems into specialized agents that interact through iterative message passing to produce unified decisions (MDAgents [12]), demonstrating promise in multi-step diagnostic reasoning [4], collaborative image interpretation [6], and even simulating hospital workflows [14]. While these systems enable complex coordination, purely generative LLMs may suffer from instability, hallucinations, and weak grounding in quantitative modality-specific signals, particularly in numerical reasoning [15]. In contrast, discriminative models provide robust, task-optimized representations from structured data. This motivates us to develop a **hybrid paradigm** (Fig 1-c) that combines the reliability of discriminative modeling with the coordination and reasoning capabilities of generative agentic systems.

### Our contribution

We propose **EPI-GUIDE**, a clinically Guideline-grounded hybrid multi-agent framework for holistic Epilepsy management, representing, to the best of our knowledge, the first exploration of agentic AI for comprehensive epilepsy care. Our framework incorporates several innovations:

– A hybrid system coupling modality-specific discriminative models with generative LLM agents, where structured discriminative predictions provide auxiliary guidance to the agentic system for final decision-making.
– A central orchestrating agent enabling structured multi-turn interactions to resolve inter-modality inconsistencies.
– Guideline grounding of the orchestrator using established international epilepsy guidelines to evaluate multimodal outputs within structured clinical pathways, enabling evidence-informed decision-making.

We evaluate our framework across two datasets spanning six epilepsy management tasks and will **publicly release a curated multi-modal, multi-task epilepsy dataset** to support future research in this direction. Our results demonstrate that combining discriminative evidence with generative coordination, particularly when augmented with clinical guidelines, leads to more reliable and comprehensive epilepsy decision-making.

## 2 Methodology

Our overall framework (Fig. 2), comprises three main components: (1) modality-specific discriminative and generative models; (2) multimodal evidence aggregation and task-driven guideline retrieval; and (3) a guideline-grounded orchestrating agent that evaluates integrated evidence for final decision-making.

**Fig. 2.**
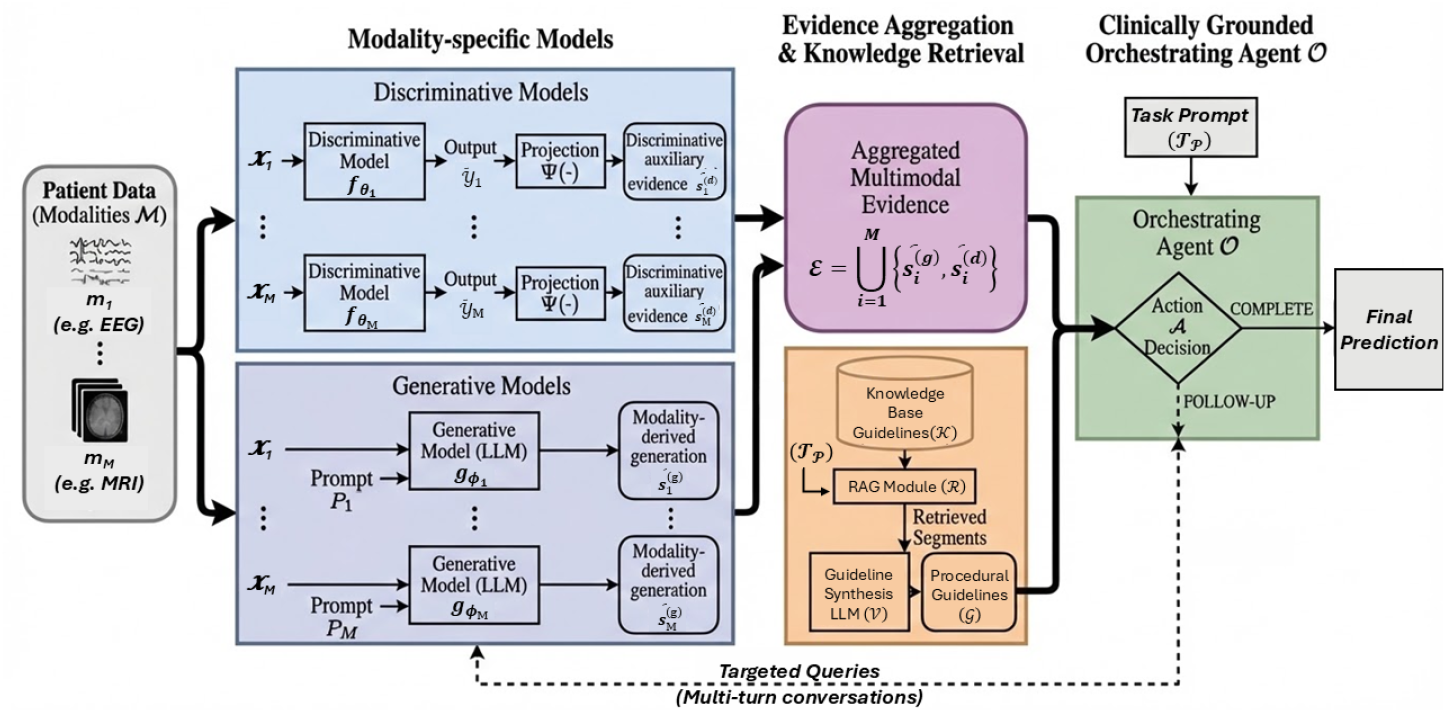
Methodology overview: EPI-GUIDE is a hybrid framework that integrates evidence from discriminative and generative models, coordinated by an epilepsy guideline–grounded orchestrating agent for multi-modal, multi-task decision-making.

### 2.1 Modality-specific Models and Evidence Aggregation

#### Clinical Practice & Multi-Modality Multi-task Problem Formulation

Epilepsy care is inherently multidisciplinary, with epileptologists, neuroradiologists, neurosurgeons, neuropsychiatrists and neuropsychologists integrating clinical history, EEG interpretation, neuroimaging findings and cognitive assessments to formulate diagnostic and treatment decisions. To emulate this workflow, let ℳ= *m*_1_, …, *m*_*M*_ denote the set of available patient modalities (e.g., EEG, MRI, …), and 𝒯= {*t*_1_, …, *t*_*T*_} the set of epilepsy management tasks (e.g., epilepsy type classification, EZ localization, …). Given inputs from ℳ, the objective is to generate predictions across 𝒯. We therefore define two complementary model families: modality-specific discriminative models and modality-specific generative models.

#### Discriminative Models

For each modality *m*_*i*_ *∈* ℳ, we define a discriminative model 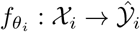, parameterized by *θ*_*i*_, where 𝒳_*i*_ denotes the modality-specific input space and 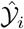 the task-specific prediction space. Each model is trained using supervised objectives appropriate to the target task *t* ∈ 𝒯 (for e.g., cross-entropy for epilepsy type classification). The resulting predictions 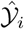 may include class probabilities, regression scores, or confidence estimates depending upon the task *t*_*i*_ ∈ 𝒯 (for e.g., classification, risk estimation, …).

To integrate with the language-based agentic framework, these structured outputs are converted into textual evidence via

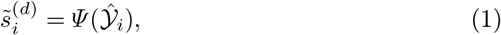

where *Ψ* (·) denotes an LLM-based transformation and 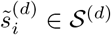 denotes modality-specific discriminative evidence used as auxiliary guidance in our framework.

#### Generative Models

In parallel, we define a generative model route through:

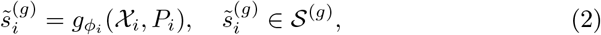

where 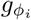 is an LLM initialized and prompted with modality-specific instructions *P*_*i*_ to produce textual clinical interpretations. Example prompt for EEG agent:

~~~
You are an EEG specialist. **Instructions:** 1) Analyze EEG montage images
for interictal discharges, ictal patterns, lateralization, …, and
2) Provide structured predictions for epilepsy type, seizure type,
… with brief clinical justification.
~~~

Finally, the discriminative 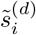 and generative 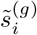 outputs are then aggregated as

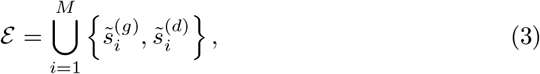

forming the unified multimodal evidence passed to the orchestrating agent.

### 2.2 Clinically Grounded Orchestrating Agent

Epilepsy management follows structured diagnostic and treatment pathways defined by international guidelines and multidisciplinary consensus. Motivated by this workflow, we introduce a guideline-grounded orchestrating agent built on Retrieval-Augmented Generation (RAG) to analyze the aggregated evidence.

Our knowledge base 𝒦 is **curated by expert neurologists** and comprises documents on epilepsy guidelines from the International League Against Epilepsy (ILAE) [26,1], National Institute for Health and Care Excellence (NICE) [19], and epilepsy textbooks, covering epilepsy type classification, drug-resistant epilepsy, ASM selection, surgical candidacy, and prognosis. The corpus is indexed into a vector database and segmented into semantic chunks. Given a task query 𝒯_*p*_, the RAG module ℛ retrieves relevant guideline segments, which are synthesized into a procedural guideline 𝒢= 𝒱 (ℛ (𝒦)), where 𝒱 (·) denotes LLM-based guideline synthesis.

The orchestrator operates on aggregated multimodal evidence ℰ as:

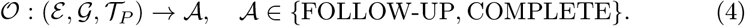

where 𝒜 is the subsequent action and 𝒯_*P*_ is a task prompt:

~~~
You are a senior epileptologist integrating multimodal reports ….
Identify concordance/discordance, resolve conflicts using guidelines,
issue FOLLOW-UP if needed, and provide final recommendations for …
~~~

If the evidence satisfies 𝒢 and is internally consistent, the action state 𝒜 is marked COMPLETE and task predictions are finalized. Otherwise, the orchestrator issues targeted FOLLOW-UP multi-turn queries to resolve discrepancies.

### 2.3 Implementation Details

#### Discriminative models

We train modality-specific discriminative predictors with weighted cross-entropy (inverse class frequency) for all classification tasks. *Clinical Text/Records:* TF–IDF (max_features = 5000, bigrams) with XGBoost, and PubMedBERT [7] fine-tuned with a lightweight classification head. *MRI/EEG:* ImageNet-pretrained ResNet-50 [8] and MedSigLIP-448 [24] (frozen vision encoder; trained head). Input images are resized to 224 2*×*24 (ResNet) or 448 *×*448 (MedSigLIP). For EEG, we additionally fine-tune an EEG foundation model (REVE-base [5]) with task-specific heads.

#### Generative models

We use MedGemma-27B-text-it [24] for clinical text and MedGemma-1.5-4B-it [24] for MRI/EEG inputs (temperature = 0.1, max tokens = 2048), each initialized with modality-specific prompts.

#### Guideline-grounded orchestrator and RAG

We use GPT-OSS-120b [21]; temperature = 0.1, tokens=2048) as the orchestrating agent. The guideline corpus is indexed with bge-base-en-v1.5 embeddings [27] and retrieved by cosine similarity w.r.t the task prompt. The orchestrator may perform up to three multi-turn rounds. All experiments were run on 8 *×*A100-80GB GPUs. Details about learning rate, optimizer, number of epochs and other hyperparameters for each individual model is available in our code repository.

## 3 Datasets

We benchmark with a standard 5-fold cross validation for two datasets.

1. **Curated Multi-modal Multi-task Epilepsy (MME) Dataset**: Our cohort was curated from publicly available PubMed papers which includes 306 epilepsy patients, with MRI available for 94 and EEG for 71 cases. This study was approved by the Alfred Hospital Ethics Committee (Ethics number 437/21). The Ethics Committee is constituted according to NHMRC guidelines and reports to the Alfred Health Executive Committee, which in turn reports to the Alfred Health Board. It contains five epilepsy management tasks, each as a three-class problem: (1) **Epilepsy type** (Focal: 216, Generalized: 30, Other: 58; *n*=304), (2) **Seizure type** (Focal Onset: 150, Generalized Onset: 57, Unknown/Other: 94; *n*=301), (3) **EZ localization** (Temporal: 151, Extratemporal: 64, Multifocal: 27; *n*=242), (4) **ASM response** (Drug-Resistant: 151, Responsive: 34, On Treatment: 57; *n*=242), and (5) **Surgical outcome** (Seizure-Free: 55, Improved: 19, No Improvement: 59; *n*=133). Due to real-world clinical variability, not all patients contain complete annotations for every task, reflecting the heterogeneous and longitudinal nature of epilepsy management.
2. **Human Intracerebral Stimulation HD-EEG Dataset [17]**: We additionally benchmark on the public simultaneous intracerebral stimulation and HD-EEG dataset [17], comprising 7 subjects and 61 stimulation sessions with paired 256-channel HD-EEG, sEEG recordings, and structural MRI. We define two tasks: (1) **EZ localization** (Temporal/Frontal/Parieto-Occipital), derived from stimulation-site coordinates; and (2) **Stimulation intensity classification** (Low ≤0.3 mA / High ≥0.5 mA).

## 4 Results

### Benchmark on MME and HD-EEG Datasets

Tables 1 and 2 compare EPI-GUIDE against modality-specific discriminative and ensemble models as well as generative baselines on both our MME and the public HD-EEG dataset [17]. On the MME cohort, generative models underperform (MedGemma-27B: 61.8% using clinical text; VLM: 29.4% using MRI), likely due to zero-shot setting; while fine-tuned discriminative models perform better (PubMedBERT: 77.6%; MedSigLIP-448: 75.4%), with the best ensemble at 78.7%, however they aren’t explainable. EPI-GUIDE achieves **85.8%** overall, outperforming PubMedBERT (+8.2), the ensemble (+7.1), and the best generative model (+24.0), with the largest gains on the data-scarce surgical outcome task (+5.7 over ensemble).

**Table 1.** Accuracy (%) on five epilepsy management tasks on our MME dataset. Results are reported as mean_*±*std_ across 5 folds or multiple evaluation rounds. T: clinical text; M: MRI; E: EEG. **Bold** - best performance, <u>underline</u> - second best.

| Method | Mod. | Epil. Type | Sz. Type | EZ Loc. | ASM Resp. | Surg. Out. | Overall |
| --- | --- | --- | --- | --- | --- | --- | --- |
| **Discriminative Models** |  |  |  |  |  |  |  |
| TF-IDF + XGBoost | T | 78.3±4.7 | <u>70.4±2.4</u> | 81.8±1.5 | 76.4±5.4 | 68.4±3.7 | 75.1±3.5 |
| PubMedBERT [7] | T | 83.9±1.7 | 65.1±4.2 | 83.5±3.6 | 75.2±2.9 | <u>80.5±4.8</u> | 77.6±3.4 |
| ResNet-50 [8] | M | 71.8±7.9 | 57.8±5.7 | 79.7±8.7 | 78.4±5.0 | 56.1±5.7 | 68.8±6.6 |
| MedSigLIP-448 [24] | M | 82.6±10.0 | 61.1±6.1 | 74.0±13.2 | 79.9±11.8 | 79.3±4.7 | 75.4±9.2 |
| ResNet-50 [8] | E | 69.1±8.4 | 56.5±11.6 | 65.3±10.4 | **87.1±6.5** | 40.7±16.9 | 63.7±10.8 |
| MedSigLIP-448 [24] | E | 74.7±13.2 | 52.2±11.8 | 73.1±13.2 | **87.1±3.2** | 75.3±12.8 | 72.5±10.8 |
| Weighted Ensemble | T+M+E | 81.9±1.2 | 69.4±1.8 | 84.3±1.5 | 78.9±1.4 | 79.0±1.1 | 78.7±1.3 |
| Stacking Ensemble | T+M+E | 82.2±1.1 | 70.1±1.7 | 83.1±1.4 | 78.5±1.3 | 75.9±1.6 | 77.9±1.4 |
| **Generative Models** |  |  |  |  |  |  |  |
| MedGemma-27B [24] | T | 72.4±4.5 | 53.2±5.1 | 67.8±4.8 | 63.2±5.3 | 52.6±6.2 | 61.8±5.2 |
| MedGemma-4B [24] | M | 52.2±5.8 | 30.0±7.2 | 56.5±6.1 | 8.1±3.4 | 0.0±0.0 | 29.4±4.5 |
| **Hybrid (Ours)** |  |  |  |  |  |  |  |
| **EPI-GUIDE** | **T+M+E** | **89.5±1.7** | **77.5±2.1** | **91.0±1.5** | <u>84.8±1.9</u> | **86.2±2.0** | **85.8±1.8** |

**Table 2.**
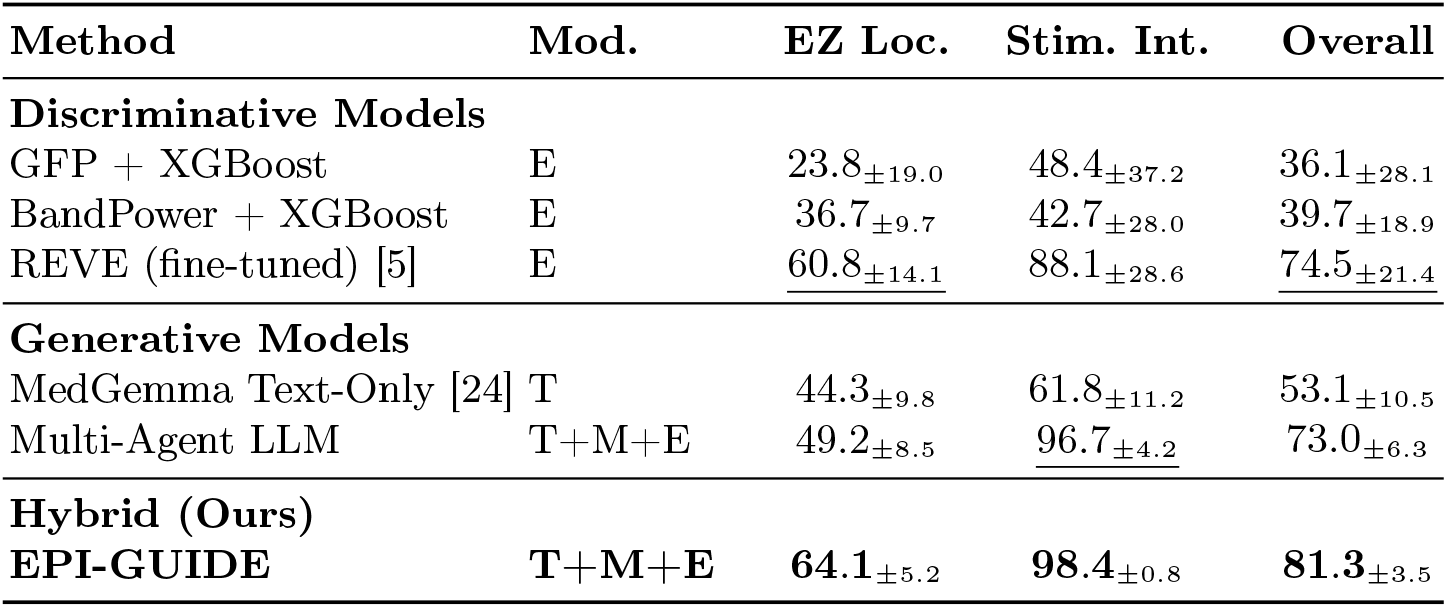
Accuracy(%) on two epilepsy management tasks on the HD-EEG dataset [17]. T: clinical text; M: MRI; E: EEG. **Bold** - best performance, underline - second best.

| Method | Mod. | EZ Loc. | Stim. Int. | Overall |
| --- | --- | --- | --- | --- |
| **Discriminative Models** |  |  |  |  |
| GFP + XGBoost | E | 23.8±19.0 | 48.4±37.2 | 36.1±28.1 |
| BandPower + XGBoost | E | 36.7±9.7 | 42.7±28.0 | 39.7±18.9 |
| REVE (fine-tuned) [5] | E | <u>60.8±14.1</u> | 88.1±28.6 | <u>74.5±21.4</u> |
| **Generative Models** |  |  |  |  |
| MedGemma Text-Only [24] | T | 44.3±9.8 | 61.8±11.2 | 53.1±10.5 |
| Multi-Agent LLM | T+M+E | 49.2±8.5 | <u>96.7±4.2</u> | 73.0±6.3 |
| **Hybrid (Ours)** |  |  |  |  |
| **EPI-GUIDE** | **T+M+E** | **64.1±5.2** | **98.4±0.8** | **81.3±3.5** |

On the public HD-EEG dataset, classical EEG features perform poorly across both tasks (36.1–39.7%). The supervised EEG foundation model REVE-base improves to 74.5% overall, with strong stimulation intensity classification (88.1%) but limited EZ region accuracy (60.8%). EPI-GUIDE achieves **81.3%** overall, surpassing REVE (+6.8) and the Multi-Agent LLM baseline (+8.3), with gains most pronounced on EZ region classification (+14.9 over Multi-Agent LLM).

### Ablation Studies for EPI-GUIDE

Table 3 evaluates each component. *(i)* Removing discriminative auxiliary guidance reduces mean accuracy by 22.9 points (85.8% → 62.9%), while removing both discriminative and guideline components collapses performance to 60.8%, matching the generative baseline. *(ii)* Disabling the guideline retrieval causes a 7.0-point drop (85.8% → 78.8%), with the largest declines on protocol-driven tasks (EZ localization and surgical outcome), demonstrating that combining discriminative and generative models with guideline grounding improves performance.

**Table 3.**
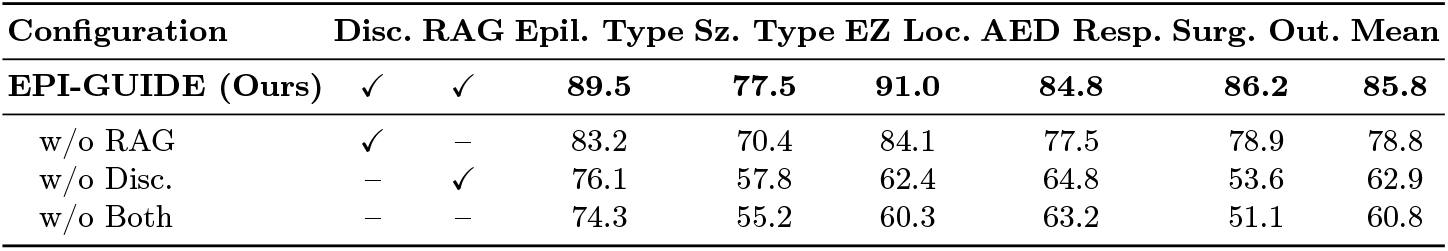
Ablation accuracy (%) of EPI-GUIDE components on MME. Disc.: discrim- inative auxiliary evidence; RAG: guideline-grounded retrieval. **Bold** indicates best.

### Qualitative Evaluation by Expert Neurologists

Fig. 3 presents a representative MME case and corresponding EPI-GUIDE outputs. We further conducted expert evaluation with neurologists on 15 cases spanning five clinical scenarios (classifier correction, multimodal fusion, surgical prediction, rare syndromes, and comprehensive classification), rated on a 1–5 Likert scale for assessing prediction reliability. EPI-GUIDE achieved 93.4% task-level accuracy (57/61) compared to 54.1% for LLM-only (33/61), and received an average rating of **3.64/5** compared to LLM-only (**2.64/5**) on reliability of predictions.

**Fig. 3.**
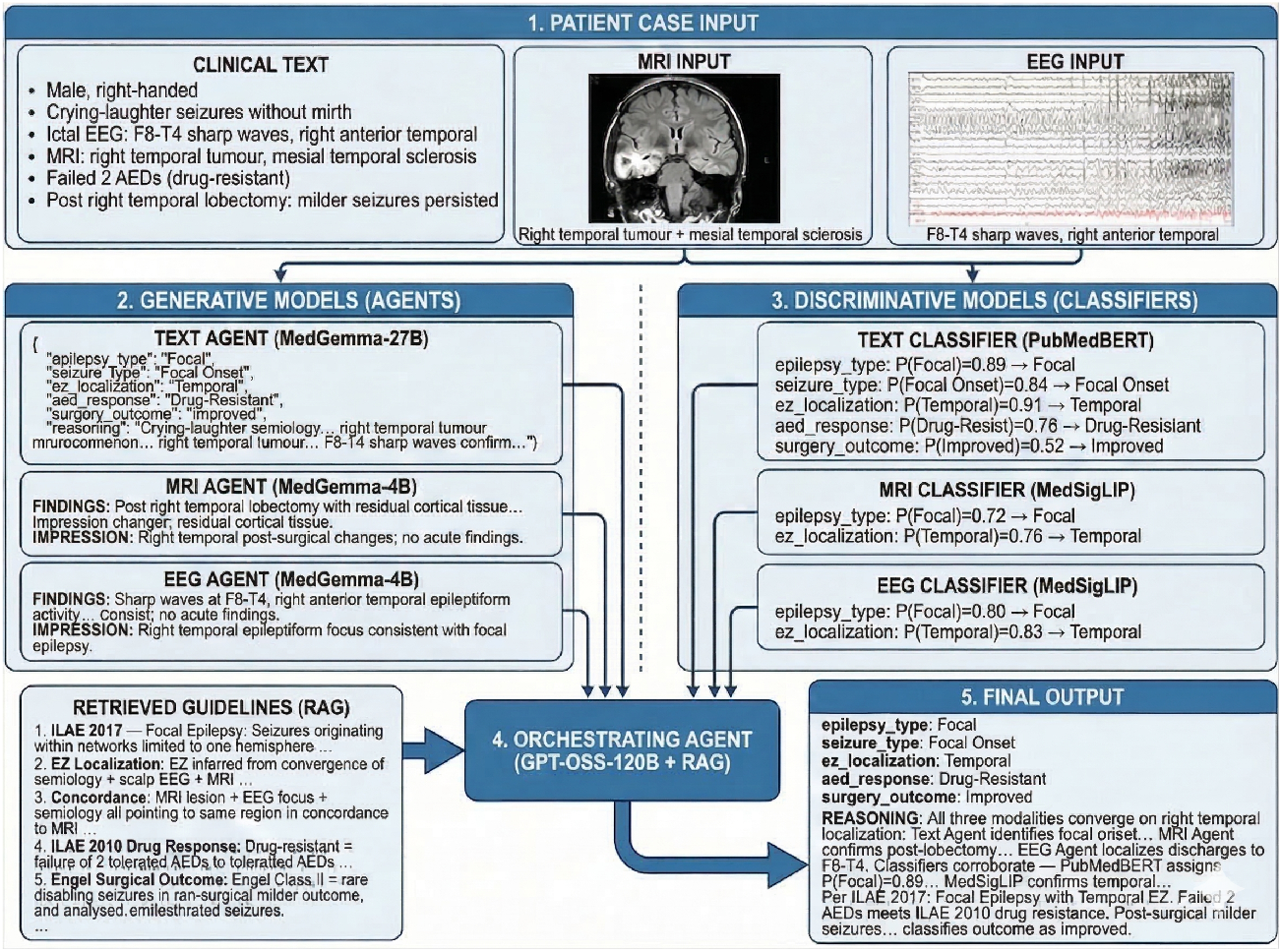
A case study demonstrating the overall flow of EPI-GUIDE.

## 5 Conclusion

In this work, we present EPI-GUIDE, to our knowledge the first exploration of agentic framework for holistic epilepsy management. Addressing the limitations of siloed, single-task models and the instability of purely generative systems, our hybrid approach integrates discriminative outputs as auxiliary evidence with guideline-grounded multi-agent coordination. EPI-GUIDE demonstrates consistent improvements across two datasets, demonstrating the promise of hybrid discriminative-generative agentic systems for robust decision-making.

## Data Availability

The data using for training model can be found at:
https://github.com/khoapham154/epi_guide.git

